# Career development awards are associated with lower career-long research attrition risk in Veterans Affairs investigators

**DOI:** 10.64898/2026.11.03.26367173

**Authors:** John J. Fraser, Samantha Hack

**Author notes:** **Corresponding Author:** John J. Fraser, DPT, PhD, FACSM, Office of Research & Development, US Department of Veterans Affairs, 810 Vermont Ave NW, Washington, DC 20420; X: @NavyPT. **Competing Interests:** Dr Fraser reports grants from Congressionally Directed Medical Research Programs, outside of the submitted work. In addition, Dr Fraser has a patent for an Adaptive and Variable Stiffness Ankle Brace (U.S. Patent 12,678,314 B2). **Individual Author Contributions:** JJF was responsible for study conception, methodological development, and data analysis. All authors contributed to the data interpretation, manuscript development, critical revision, and final approval of the study. **Disclaimer:** The authors are employees of the U.S. Government, and this work was prepared as part of their official duties. Title 17, U.S.C. §105 provides that copyright protection under this title is not available for any work of the U.S. Government. Title 17, U.S.C. §101 defines a U.S. Government work as work prepared by a military service member or employee of the U.S. Government as part of that person’s official duties. The views expressed are those of the authors and do not necessarily reflect the official policy or position of the Department of Veterans Affairs, nor the U.S. Government. The study protocol was approved by the VA Central Institutional Review Board as non-human subjects research.

## Abstract

**Objective:** To investigate research attrition and the associated factors among US Department of Veterans Affairs investigators with and without a prior Career Development Award (CDA) from fiscal year 2008 to 2026.

**Design:** Retrospective cohort study.

**Methods:** Investigators whose CDA ended between October 2008 and March 2026 were identified from the Research Analysis and Forecasting Tool (RAFT) database and compared to non-CDA peers. Extracted descriptive variables included sex, investigator type, doctoral degree, salary source, specialty, and research area. Attrition, derived from the electronic Project Management and Information System, was defined when an investigator ceased serving as a principal or coinvestigator on VA-funded research. Analysis utilized attrition percentages, crude count ratios (CR), chi-squared statistics, and multivariable binomial regression to assess adjusted odds ratios (ORadj).

**Results:** Investigators with a prior CDA were 42% less likely to attrite from VA research engagement compared to non-CDA counterparts (CR 95% CI, 0.50-0.67, p<.001). They maintained high engagement regardless of demographic or professional variables (engagement ranges: 66.7%-93.8%). Clinician-scientists were 34% less likely to attrite from science (OR_adj_ 0.54) compared to non-clinician scientists. The 1.68-times higher crude attrition risk for non-VA salaried investigators became non-significant after adjusting for other factors. Attrition was significantly higher in basic science and engineering (OR_adj_ 3.90), endocrinology, metabolism, and nutrition (OR_adj_ 4.24), infectious disease (OR_adj_ 3.80), aging and geriatrics (OR_adj_ 3.56), and cardiopulmonary and vascular research (OR_adj_ 2.70) compared to health services and epidemiology.

**Conclusion:** Prior CDA recipients demonstrate significantly lower research attrition in VA research.

## INTRODUCTION

The US Department of Veterans Affairs (VA) Office of Research and Development (ORD) developed the intramural Career Development Award (CDA) program in 1990 as a way to develop early career investigators and to build capability in the VA research enterprise.^1^ The CDA is a powerful funding mechanism that provides salary support and project funds to support mentored research and continued training to develop early-career investigators.^2^ The CDA-2 (IK2) award provides up to five years of support. Clinician-scientists are eligible for application for this award within 10 years of graduation from their clinical degree, with non-clinician scientists eligible within five years of graduation from their PhD or equivalent terminal degree.^2^ As a result of this funding, early-career investigators further develop their knowledge, skills, and abilities needed to be competitive when competing for merit-review level (I01) awards.^1^

In our previous investigation of the CDA program in rehabilitation scientists, we found that the protected research time and experiential learning received during this training award provided a career-long competitive advantage for obtaining subsequent merit-review awards.^1^ In addition to the increased technical expertise garnered from the CDA, we posited that this time also provided the investigator an opportunity to develop a professional reputation within the organization and time to develop research questions salient to Veteran populations and in alignment with VA organizational priorities.^1^ With the increased odds of award funding and specialized expertise in Veterans’ health, it is plausible that the CDA influenced career-long engagement in VA research and the characteristics of the investigators.

While the CDA is likely very important factors that may have an influence on career-long engagement in VA research, there are additional factors that may influence this outcome that warrant investigation. Sex,^3^ investigator type (clinician vs non-clinician),^4^ doctoral degree type,^5,6^ specialty,^7^ area of research,^8^ and salary source^9^ have been shown to impact funding success and career trajectory in analyses of other academic populations. Therefore, empirical evaluation of the subpopulations of scientists with the greatest attrition from VA research may provide insight regarding opportunities for organization improvement to optimize the return on investment for these investigators. The purpose of this research was to investigate the count ratio and associated factors of research attrition in VA investigators with and without a prior CDA. We hypothesized that investigators with a CDA would demonstrate lower research attrition compared to their counterparts who did not benefit from this mentored research award.

## METHODS

A retrospective cohort study of research attrition in all VA investigators with and without a history of completion of a CDA-2 (IK2) from fiscal year 2008 to 2026 was performed. The factors of sex, investigator type (clinician vs non-clinician), doctoral degree type, salary source, specialty, and research area were evaluated. This study was determined to be non-human subjects research by the VA Central Institutional Review Board. The Strengthening the Reporting of Observational Studies in Epidemiology (STROBE) guidelines were used to guide reporting.^10^

Investigators with a CDA which ended between October 2008 and March 2026 were identified from the Research Analysis and Forecasting Tool (RAFT) database (VA Office of Research and Development, Washington, DC), an internal platform used to manage research funds and track expenditures. Descriptive variables of sex, investigator type (clinician vs non-clinician), doctoral degree type (academic doctorate, clinical doctorate, dual doctorate), salary source (Medical Research Program, Health Services Research Program, VA funding outside of ORD, not salaried by the VA), specialty, and research area were extracted from RAFT in April 2026. Research attrition was operationally defined in this study when a VA investigator was listed as inactive in the electronic Project Management and Information System, an indicator triggered when the investigator no longer served as a principal investigator or coinvestigator on any VA funded research. This status did not preclude continued employment in the VA in administrative or clinical capacities, only the cessation of VA-funded research.

Research attrition percentages, crude count ratios (CR), 95% confidence intervals (CIs), risk differences, attributable risk, and chi-squared statistics were calculated using Microsoft® Excel for Mac (version 16.98, Microsoft Corp., Redmond, WA) and a custom epidemiological spreadsheet.^11^ A multivariable binomial regression model was employed to assess the adjusted odds ratios (OR_adj_) on attrition on R (version 4.6, The R Foundation for Statistical Computing, Vienna, Austria). The level of significance was *p* < 0.05 for all analyses. CR point estimates were considered statistically significant if CIs did not cross the 1.00 threshold. Convergence of p-values, effect sizes, and 95% confidence intervals were considered when evaluating significant findings.

## RESULTS

VA investigators with a prior CDA were 42% less likely to attrite from VA research engagement over the course of a career compared to their counterparts without a prior CDA (**Table 1**). Investigators with a prior CDA demonstrated a high degree of engagement in VA-funded research regardless of sex (range 81.1%-81.6%), investigator type (range 76.8%-84.8%), degree type (range 78.9%-84.5%), salary source (66.7%-81.8%), specialty (range 66.7%-93.8%), and research area (range 67.3%-90.0%) (**Table 2**). **Table 3** details the results of the multivariable logistic regression evaluating factors associated with research attrition among VA investigators with a prior CDA. In the evaluation of research attrition risk, non-clinician scientists were 1.51 more likely (85% greater adjusted odds) of disengagement from VA research compared to their clinician-scientist counterparts. Investigators who were not salaried by the VA were 1.68 times more likely to attrite compared those funded by the ORD Medical Research Program, but this effect washed out and was non-significant once adjusted for the other factors. Investigators who conducted basic science & engineering (CR: 3.27: OR_adj_: 3.90), cardiopulmonary and vascular (CR was not significant, OR_adj_: 2.70), endocrinology, metabolism, and nutrition (CR: 3.00, OR_adj_: 4.24), aging and geriatrics (CR: 2.93, OR_adj_: 3.56), and infection disease (CR: 2.61, OR_adj_: 3.80) research were significantly more likely to attrite than their health services and epidemiology investigator counterparts. While crude CR for otolaryngology, ophthalmology, and speech was statistically significant (2.63), this effect was not significant in the multivariable analysis once the other variables were factored. There were no other significant findings.

**Table 1.** Descriptive statistics and career-long research attrition risk in investigators with and without a prior Veterans Affairs Career Development Award (CDA).

|  | Research Engaged |  | Attrited |  | CR | 95% CI |  | RD% | AR% | p-value |
|  | Count | % | Count | % |  | LL | UL |  |  |  |
|---|---|---|---|---|---|---|---|---|---|---|
| Investigators with a CDA | 679 | 81.3 | 156 | 18.7 | **0.58** | **0.50** | **0.67** | **-13.7** | **-73.4** | **<.001** |
| Investigators without a CDA | 2239 | 67.9 | 1061 | 32.2 | *Reference* |  |  |  |  |  |

CR, count ratio; CI, confidence interval; LL, lower limit; UL, upper limit; RD, risk difference; AR, attributable risk. Bolded values indicate statistical significance

**Table 2.** Descriptive statistics of investigators with prior Veterans Affairs (VA) Career Development Award and crude risk calculations of career-long research attrition in the US Department of Veterans Affairs.

|  |  | Research Engaged |  | Attrited |  | CR | 95% CI | RD% | AR% | p-value |
|---|---|---|---|---|---|---|---|---|---|---|
|  |  | Count | % | Count | % |  |  |  |  |  |
| Sex: | Male | 315 | 81.6 | 71 | 18.4 | *Reference* |  |  |  |  |
|  | Female | 364 | 81.1 | 85 | 18.9 | 1.03 | 0.77-1.37 | 0.5 | 2.8 | .84 |
| Investigator Type: | Non-clinician | 282 | 76.8 | 85 | 23.2 | Reference |  |  |  |  |
|  | Clinician | 397 | 84.8 | 71 | 15.2 | **0.66** | **0.49-0.87** | **-8.0** | **-52.7** | **.003** |
| Degree: | Academic doctorate | 366 | 78.9 | 98 | 21.1 | *Reference* |  |  |  |  |
|  | Clinical doctorate | 238 | 84.1 | 45 | 15.9 | 0.75 | 0.55-1.04 | -5.2 | -32.8 | .08 |
|  | Dual doctorate (e.g. MD PhD) | 71 | 84.5 | 13 | 15.5 | 0.73 | 0.43-1.24 | -5.6 | -36.5 | .24 |
| Salary Source: |  |  |  |  |  |  |  |  |  |  |
|  | Medical Research Program | 294 | 80.1 | 73 | 19.9 | *Reference* |  |  |  |  |
|  | Health Services Research Program | 9 | 81.8 | 2 | 18.2 | 0.91 | 0.26-3.26 | -1.7 | -9.4 | * |
|  | VA, external to ORD | 338 | 84.5 | 62 | 15.5 | 0.78 | 0.57-1.06 | -4.4 | -28.3 | .11 |
|  | Not salaried by the VA | 38 | 66.7 | 19 | 33.3 | **1.68** | **1.10-2.55** | **13.4** | **40.3** | **.02** |
| Specialty: |  |  |  |  |  |  |  |  |  |  |
|  | Public Health & Social Sciences | 55 | 83.3 | 11 | 16.7 | *Reference* |  |  |  |  |
|  | Basic Sciences & Engineering | 57 | 73.1 | 21 | 26.9 | 1.62 | 0.84-3.10 | 10.2 | 38.1 | .14 |
|  | Allied Health & Nursing | 23 | 67.6 | 11 | 32.4 | 1.94 | 0.94-4.01 | 15.7 | 48.5 | .07 |
|  | Family/Internal Medicine & Subspecialties | 191 | 83.4 | 38 | 16.6 | 1.00 | 0.54-1.84 | -0.1 | -0.4 | .99 |
|  | Geriatrics and Aging | 18 | 90.0 | 2 | 10.0 | 0.60 | 0.14-2.49 | -6.7 | -66.7 | * |
|  | Mental Health | 196 | 83.8 | 38 | 16.2 | 0.97 | 0.53-1.80 | -0.0 | -2.6 | .93 |
|  | Neurology & Neuroscience | 39 | 79.6 | 10 | 20.4 | 1.22 | 0.57-2.65 | 3.7 | 18.3 | .61 |
|  | Oncology | 15 | 93.8 | 1 | 6.2 | 0.38 | 0.05-2.70 | -10.4 | -166.7 | * |
|  | Pathology & Laboratory Medicine | 8 | 66.7 | 4 | 33.3 | 2.00 | 0.76-5.25 | 16.7 | 50.0 | * |
|  | Pharmacy & Pharmacology | 7 | 70.0 | 3 | 30.0 | 1.80 | 0.61-5.35 | 13.3 | 44.4 | * |
|  | Physical Medicine & Rehabilitation | 25 | 86.2 | 4 | 13.8 | 0.83 | 0.29-2.38 | -2.9 | -20.8 | * |
|  | Surgery & Subspecialties | 26 | 74.3 | 9 | 25.7 | 1.54 | 0.71-3.37 | 9.1 | 35.2 | .28 |
|  | Other Medical Specialties | 18 | 85.7 | 3 | 14.3 | 0.86 | 0.26-2.78 | -2.4 | -16.7 | .80 |
| Research Area: |  |  |  |  |  |  |  |  |  |  |
|  | Health Services & Epidemiology | 99 | 90.0 | 11 | 10.0 | *Reference* |  |  |  |  |
|  | Basic Sciences & Engineering | 33 | 67.3 | 16 | 32.7 | **3.27** | **1.64-6.51** | **22.7** | **69.4** | **<.001** |
|  | Cardiopulmonary & Vascular | 64 | 81.0 | 15 | 19.0 | 1.90 | 0.92-3.91 | 9.0 | 47.3 | .08 |
|  | Endocrinology, Metabolism, & Nutrition | 14 | 70.0 | 6 | 30.0 | **3.00** | **1.25-7.18** | **20.0** | **66.7** | **.02** |
|  | Otolaryngology, Ophthalmology, & Speech | 14 | 73.7 | 5 | 26.3 | **2.63** | **1.03-6.73** | **16.3** | **62.0** | **.05** |
|  | Gastroenterology | 20 | 83.3 | 4 | 16.7 | 1.67 | 0.58-4.79 | 6.7 | 40.0 | .35 |
|  | Genitourinary | 21 | 77.8 | 6 | 22.2 | 2.22 | 0.90-5.47 | 12.2 | 55.0 | .08 |
|  | Aging & Geriatrics | 29 | 70.7 | 12 | 29.3 | **2.93** | **1.40-6.11** | **19.3** | **65.8** | **.003** |
|  | Immunology & Rheumatology | 18 | 78.3 | 5 | 21.7 | 2.17 | 0.83-5.66 | 11.7 | 54.0 | .12 |
|  | Infectious Diseases | 17 | 73.9 | 6 | 26.1 | **2.61** | **1.07-6.34** | **16.1** | **61.7** | **.04** |
|  | Mental Health & Behavioral Sciences | 164 | 85.9 | 27 | 14.1 | 1.41 | 0.73-2.74 | 4.1 | 29.3 | .30 |
|  | Neurology, Neuroscience & Sleep | 58 | 81.7 | 13 | 18.3 | 1.83 | 0.87-3.86 | 8.3 | 45.4 | .11 |
|  | Oncology & Hematology | 29 | 80.6 | 7 | 19.4 | 1.94 | 0.81-4.64 | 9.4 | 48.6 | .14 |
|  | Rehabilitation & Biomechanics | 41 | 83.7 | 8 | 16.3 | 1.63 | 0.70-3.81 | 6.3 | 38.8 | .26 |
|  | Substance Abuse | 22 | 78.6 | 6 | 21.4 | 2.14 | 0.87-5.29 | 11.4 | 53.3 | .10 |
|  | Other | 35 | 81.4 | 8 | 18.6 | 1.86 | 0.80-4.31 | 8.6 | 46.3 | .15 |

CR, count ratio; RD, risk difference; AR, attributable risk; ORD, Office of Research & Development. Bolded values indicate statistical significance. \* X<sup>2</sup> statistic not calculated due to counts less than 5.

**Table 3.**
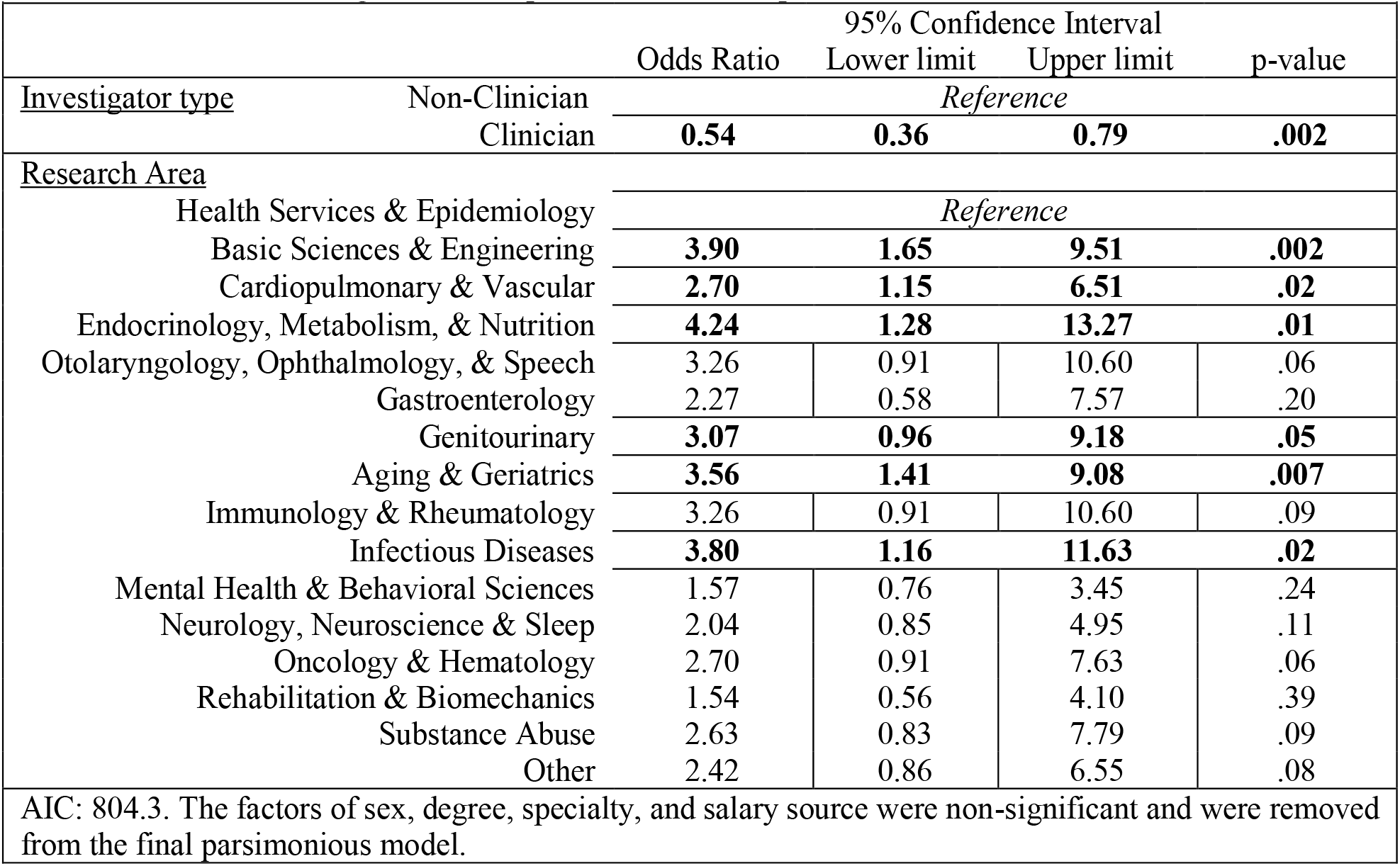
Results of the multivariable logistic regression evaluating associated factors with research attrition in Veterans Affairs investigators with a prior Career Development Award.

## DISCUSSION

The primary finding of this study was that scientists with a prior CDA demonstrated a high degree of continued engagement in research throughout their VA careers, with evidence of significantly lower research attrition compared to their counterparts without a prior CDA. While there were no significant differences in continued engagement in VA research attributable to investigators’ sex, degree type, and specialty, there were significantly higher rates of research attrition in non-clinician scientists and those engaged in the basic, engineering, and internal medicine scientific areas of study. This study is the first to the authors’ knowledge to have evaluated the associated factors of engagement in research in VA scientists throughout a career.

VA investigators with a prior CDA demonstrated significantly lower research attrition compared to their counterparts without a prior CDA. One explanation for this finding may be related to the increased merit-review award funding success rates of prior CDAs. In a study evaluating the effects of the CDA on subsequent merit review funding awards, rehabilitation scientists with a prior CDA were found to have an adjusted 1.54 greater odds of funding compared to their counterparts who did not have this developmental experience.^1^ This may be evidence of the “Matthew Effect,” a phenomenon where accumulation of resources and recognition in early career scientists results in increased subsequent scientific success.^12,13^ The increased subsequent scientific success in VA investigators CDAs may motivate continued application and execution of research throughout a career in the VA. When considering the system necessitates procurement of soft funding for continued functioning as a scientific investigator, lower funding success likely drives career decision-making, motivating or necessitating some investigators to seek funding outside of the VA, or a functional shift to more administrative, academic teaching, or clinical roles.

Among VA investigators with a prior CDA, clinician-scientists demonstrated 85% increased odds when compared to their non-clinical counterparts in the adjusted analysis of continued VA research engagement over the course of a career. These findings are likely related to the career flexibility provided to licensed and credentialed VA clinician-scientists, who comprise approximately 60% of the scientific workforce.^14^ During periods of research funding dearth, clinician-scientists in the VA have the ability to increase the number of patient care hours needed to cover their salaries. When they successfully apply for research funding, they can adjust their clinical effort once again as they pivot back to increasing research roles. Non-clinician-scientists do not have this flexibility and may need to take on administrative responsibilities within the VA, increase their effort in the academic affiliate and reduce the number of hours in VA, or leave the VA outright for other pursuits. With the ebb and flow of research funding, policies or bridge funding that provide non-clinician investigators with the same opportunity to pivot within the VA ecosystem during research funding gaps may promote greater career-long engagement in research. While competitive salary awards such as the VA Research Career Scientist (RCS, IK6) award, funding that is reserved for established scientists who have a demonstrated body of impactful work and VA service, provide salary support and stability between study awards for non-clinician scientists, there are only 115 out of approximately 1480 active non-clinician investigators (at the time of writing in September 2026) that hold these awards. Another potential factor for the high engagement of clinician-scientists throughout a career is job satisfaction. In a study evaluating VA physicians, clinician-scientists demonstrated a 23% increased adjusted odds in overall job satisfaction when compared to their colleagues who have only clinical roles^15^. There is unfortunately no corresponding research regarding job satisfaction in VA non-clinician scientists, which represents an evidence gap and opportunity for future research funding.

VA investigators in the basic sciences, engineering, and internal medicine research areas demonstrated the greatest odds of research attrition compared to investigators engaged in health services and epidemiological research. A plausible reason for this observation is the recruitment of investigators in these research areas to industry, who may provide more lucrative and stable opportunities in the device and pharmaceutical developmental space. In order to maintain expertise in these areas and when scientific workforce-shaping dictates, one method to counter attrition in these research areas is to provide increased and prioritized funding opportunities to incentivize continued VA research and mitigate attrition. Another approach to balancing attrition in these critical areas is to find replacements through targeted and incentivizing recruitment and growth through the CDA program. Shifting from the federal general schedule (GS) to a sliding pay scale for VA scientists may also provide some flexibility toward increasing pay of scientists in critical areas, mitigating the federal to private sector pay differential, and help to optimize retention. For scientists and engineers in the development space, increasing the percentage of royalties paid through licensing intellectual property could also incentivize retention. Increased investment in the RCS program for non-clinician scientists performing in critical research areas may help to encourage continued service and research execution in the VA.

In addition to these known variables which impact researcher retention, new factors are emerging. In a 2025 survey of 1,329 investigators with National Institutes of Health (NIH) K awards, the NIH equivalent to a VA CDA, 70% of respondents reported they were somewhat less to much less likely to continue conducting research due to federal policy changes.^16^ Future analyses should consider how perceptions of research career stability and potential may impact research career attrition in the VA.

Title 38 U.S.C. Section 7301-7303 tasks the Veterans Health Administration with the medical care and treatment of Veterans, education and training of health care professionals, and execution of a program of diverse medical research. This integrated health care mission in service of the highest quality of care for U.S. Veterans and the ability to see one’s work translated into clinical care has long been an incentive for researchers to pursue VA careers.

However, as evidenced by these findings, there are factors which can sustain this commitment over the career course. Some have already been implemented, such as reciprocal investment in researchers’ training and development through Career Development Awards. Others should be explored with input from both clinician and non-clinician scientists to address profession specific causes of attrition such as lack of flexible funding streams, private-public sector income disparities, and federal policy uncertainty. A proactive approach to researcher retention will ensure innovative, high-quality health care for Veterans supported by career VA scientists.

There are limitations to this study. First, this study relied on coding of primary research area and discipline in the RAFT database, which data is entered by the scientific program managers following notice of award. While it is plausible that coding heterogeneity may have biased results, comfort may be taken that there is training in ORD to increase calibration and reduce coding variability. It is plausible that variability in factors such as the size of the scientific program, resources available, and the enthusiasm and support of VA medical center executive leadership toward science may have influenced the findings in the current study. Lastly, there are likely other personal factors that may have influenced the decision to pivot out of VA research. Future research regarding the intrinsic, sociocultural, and environmental facilitators and barriers to research that may influence the decision to engage in research in the government is warranted.

## CONCLUSION

VA scientists with a prior CDA demonstrated a high degree of continued engagement in research throughout their careers, with evidence of significantly lower research attrition compared to their counterparts without a prior CDA. While there no significant differences in continued engagement in VA research in regardless of the investigators’ sex, degree type, and specialty, there were significantly higher rates of research attrition in non-clinician scientists and those engaged in the basic, engineering, and internal medicine scientific areas of study.

## Data Availability

Restrictions apply to the availability of these government datasets. Data may be requested directly from the US Department of Veterans Affairs, subject to official application and approval.

## Acknowledgements

Paul Karanam and Waheed Sheikh for their assistance in data extraction.

## Data Availability

The datasets generated and/or analyzed during the current study are not publicly available due to privacy regulations, but they may be made available on reasonable request.

